# Blood-brain barrier integrity decreases with higher blood pressure, a 7T DCE-MRI study

**DOI:** 10.1101/2024.01.02.24300739

**Authors:** Marieke van den Kerkhof, Joost J.A. de Jong, Paulien H.M. Voorter, Alida Postma, Abraham A. Kroon, Robert J. van Oostenbrugge, Jacobus F.A. Jansen, Walter H. Backes

## Abstract

**Background:** Blood-brain barrier (BBB) integrity is presumed to be impaired in hypertension, resulting from cerebral endothelial dysfunction. Hypertension antedates various cerebrovascular diseases, such as cerebral small vessel disease (cSVD), and is a risk factor for developing neurodegenerative diseases for which BBB disruption is a preceding pathophysiological process. In this study, we investigated the relation between hypertension, current blood pressure and BBB leakage in human subjects.

**Methods:** BBB leakage was determined in twenty-two patients with hypertension and nineteen normotensive controls, age- and sex-matched (median age[range]:65[45-80] years,19 males), using a sparsely time-sampled contrast-enhanced 7 Tesla MRI protocol. Structural cSVD markers were visually rated. Multivariable regression analyses, adjusted for age, sex, cardiovascular risk factors and cSVD markers, were performed to determine the relation between hypertension status, systolic and diastolic blood pressure (SBP and DBP), mean arterial pressure (MAP), drug treatment, and BBB leakage.

**Results:** Both hypertensive and normotensive participants showed mild scores of cSVD. BBB leakage did not differ between hypertensive and normotensive participants, however was significantly higher for SBP, DBP and MAP in the cortex, and DBP and MAP in the grey matter. Effectively treated patients showed less BBB leakage than those with current hypertension.

**Conclusion:** BBB integrity in the total and cortical grey matter decreases with increasing blood pressure, but is not related to hypertension status. These findings show that BBB disruption already occurs with increasing blood pressure, before the presence of overt cerebral tissue damage. Additionally, our results suggest that effective antihypertensive medication has a protective effect on the BBB.

Registered at: https://trialsearch.who.int/; Unique identifier: NL7537

## Introduction

Hypertension is a highly prevalent condition, and its incidence increases in an aging population.^1^ In addition to the heart and kidneys, the brain is highly susceptible to high blood pressure, which consequently may result in neurological diseases, such as stroke and dementia.^2^ Hypertension causes vascular remodelling, which includes both structural and functional alterations of the vessel walls. Over time, these changes lead to endothelial dysfunction and to breakdown of the blood-brain barrier (BBB), with a detrimental effect on the neuronal microenvironment and brain health.^3,4^

The BBB is a highly specialized structure in the cerebral vessel walls and preserves the biological homeostasis of the central nervous system through selective supply of nutrients to the brain tissue and removal of waste products.^5^ Loss of BBB integrity increases the permeability of this barrier, and thereby allows neurotoxins to accumulate in the brain tissue. Although hypertension is assumed to have a negative effect on BBB integrity, the precise pathophysiology has not yet been elucidated. Potentially, vascular oxidative stress and inflammation lead to dysfunction of endothelial cells and degeneration of pericytes, consequently resulting in a disrupted BBB.^3^ A large number of studies have investigated BBB impairment and hypertension in animal studies and found an increased permeability in several regions for hypertensive models.^6^ Others reported subtle BBB impairment in human patients with disorders such as preeclampsia, cerebral small vessel disease (cSVD), and dementia.^7–10^ While it is often assumed that hypertension leads to BBB disruption, the relation of hypertension or high blood pressure and BBB permeability has not yet been investigated explicitly in human subjects.

The most commonly used technique to measure BBB integrity is dynamic contrast-enhanced (DCE) MRI. This method uses the intravenous administration of a paramagnetic contrast agent, which is subsequently followed over time by acquiring a continuous series of MRI scans after contrast administration. The leakage of the contrast agent from the blood circulation to the brain tissue is quantified by the signal intensity changes, and converting these temporal changes to blood concentrations. Recently, we proposed a highly sensitive DCE-method at 7 Tesla, where instead of the commonly continuously acquired T_1_-weighted images, only two quantitative post-contrast T_1_-maps were acquired.^11^

In the current study, we investigated the relation between hypertension, blood pressure and BBB leakage in human subjects. Therefore, we applied the sparsely time-sampled, dynamic contrast-enhanced MRI protocol to patients with essential hypertension and normotensive, healthy controls, to investigate group differences in BBB leakage rates.

## Methods

### Data availability

The data that support the findings of this study are available from the corresponding author upon reasonable request.

### Study population

Between July 2019 and July 2021, 23 patients with essential hypertension and 20 normotensive, healthy controls were included, matched on age and sex. The patients with essential hypertension were recruited from the outpatient internal medicine clinic of the Maastricht University Medical Center, The Netherlands, and through a recruitment website (hersenonderzoek.nl). Essential hypertension was defined as a mean blood pressure of ≥ 135 mmHg systolic or ≥ 85 mmHg diastolic, or both, when measured for 30 minutes with an automated blood pressure monitor. Additionally, participants who took antihypertensive medication were also defined as hypertensive. The recruitment of healthy controls took place via advertisements in the local newspaper, in the hospital, and on a recruitment website (hersenonderzoek.nl). Inclusion criteria for all participants were: age between 30-90 years old and eligibility to undergo 7 Tesla MRI with contrast agent administration. Exclusion criteria were history of secondary hypertension, diabetes mellitus, ischemic heart disease, hemorrhagic stroke, or preeclampsia, and no diagnosis of obstructive sleep apnea syndrome, body mass index (BMI) > 32 kg/m^2^, and contraindications for the gadolinium-containing contrast agent, including glomerular filtration rate (eGFR) <30 mL/min.

Before study participation, all participants gave written informed consent. The study was approved by the local Medical Ethical Committee of Maastricht University Medical Center (Trial NL7537).

### Demographics

Demographics were recorded, which included age, sex, type of antihypertensive medication (if applicable), other types of medication (if applicable), BMI, smoking status (current and history) and alcohol use.

### Blood pressure measurement

All participants underwent automated blood pressure measurement (Dinamap, GE Healthcare, Chicago, USA) preceding the MRI-scan, which was performed on the same day. This measurement was performed by the same trained investigator for all participants (MvdK). First, the blood pressure was consecutively determined at both arms with a single measurement. The arm which acquired the highest blood pressure was used for the subsequent measurements every 5 minutes during 30-45 minutes. Throughout this time, the participant was kept seated solitary in the room, and was instructed to stay awake. To obtain the blood pressure characteristics, the last five measurements were averaged and used in the data analysis, yielding systolic and diastolic blood pressure (SBP and DBP, respectively), and mean arterial pressure (MAP).

### MRI

Brain images were acquired with a 7 Tesla MRI (Magnetom, Siemens Healthineers, Erlangen, Germany) using a 32-channel phased-array head coil. Dielectric pads were placed on both sides of the participant’s neck, proximal to the temporal lobes, for improvement of B1+ field homogeneity across the brain. Anatomical and DCE-images were acquired. The DCE-protocol consisted of quantitative precontrast 3D T1 mapping using a Magnetization-Prepared 2 Rapid Acquisition Gradient Echo (MP2RAGE) sequence (TR/TE = 5000/2.47 ms, TI_1_/TI_2_ = 2700/900 ms, cubic voxel size = 0.7 mm, acquisition time = 8:00 min:s), followed by a 3D fast gradient echo T1-weighted perfusion (volumetric interpolated brain examination, VIBE) sequence (TR/TE = 3.7/1.3 ms, cubic voxel size = 2.0 mm, acquisition time = 2:47 min:s). After the first three volumes were acquired, the contrast agent (1.0 molar Gadobutrol, 3 mL for each participant) was injected with an infusion rate of 0.3 mL/s followed by a saline flush (20 mL). Finally, two postcontrast T1 maps (TR/TE = 4000/2.32 ms, TI_1_/TI_2_ = 2700/900 ms, cubic voxel size = 1.2 mm, acquisition time = 4:16 min:s) were acquired, of which the first postcontrast T1 map was acquired immediately after the dynamic perfusion scan series and the last T1 map was acquired approximately 25 minutes after the start of CA injection. More details about the scan parameters and this protocol are reported in Table S1 and Kerkhof et al. (2021).^11^ For optimal segmentation of grey matter (GM) and white matter (WM), a T2-weighted fluid–attenuated inversion recovery (FLAIR) sequence (TR/TE/TI = 8000/303/2330 ms; cubic voxel size: 1 mm, acquisition time: 6:59 min:s) was acquired in between the two postcontrast T1-maps. Additionally, for the rating of microbleeds and perivascular spaces (PVS), a susceptibility weighted imaging (SWI) sequence was acquired before contrast administration, and T2-weighted images were acquired after contrast administration, respectively.

### Image analysis

Preprocessing of the quantitative T1 maps consisted of bias field correction, skull stripping and gradient distortion correction. To correct for head displacements, all acquired images were spatially registered to the same reference image, which was the first postcontrast T1 map. Subsequently, GM and WM were automatically segmented using the precontrast T1 and FLAIR images as input (FreeSurfer version 6.0.5).^12^ The segmentations were visually verified, and manually corrected when required. This resulted in four tissue regions of interest (ROI): total GM and total WM, cortical and deep GM). The subject-specific vascular input function (VIF) was derived from an ROI manually delineated in the superior sagittal sinus.

The leakage maps were determined by applying the graphical Patlak approach to the concentration time-courses of each brain tissue voxel and the VIF, resulting in a Patlak plot, in which the slope represents the BBB leakage rate, *K_i_* [min^−1^], and the intercept yields the blood plasma volume fraction, *v_p_* [−].^13^ Per ROI, the mean *K_i_* and *v_p_* of all voxels within each ROI were calculated and used as physiological measures. Outlier correction was performed by considering the *K_i_* and *v_p_* values in these ROIs located in the 95% confidence interval.^11^ All data analyses were performed using custom-made code in the Matlab programming environment (2019b, 9.2.0; Mathworks, Nattick, MA, USA).

### cSVD rating

Four cSVD-markers, i.e., white matter hyperintensities (WMH), lacunar infarcts, microbleeds and MRI-visible PVS, were visually rated by one trained neuroscientist (MvdK: >2 years of experience) under supervision of an experienced neuroradiologist (AAP: >20 years of experience) and were combined into a global cSVD-score per participant.^14^ The WMH load was rated using the Fazekas scale.^15^ When (early) confluent deep white matter hyperintensities were present (Fazekas score 2 and 3) or irregular periventricular hyperintensities which extended into the deep white matter (Fazekas score 3), one point was assigned to the global cSVD-score.^14^ Furthermore, one point was assigned when 1 or more asymptomatic lesions, i.e., lacunar infarcts, were present.^16^ Another point was assigned when 1 or more microbleeds were present in the basal ganglia, since these were associated with hypertension.^16^ Lastly, PVS were rated in the basal ganglia, as these have also been shown to be associated with hypertension.^17,18^ The PVS score was established in the slice with the highest number of PVS, and graded in four groups: 0: ≤ 10, 1: 11-25, 2: 26-40, 3: ≥ 41 PVS, as previously published.^19^ A last point was assigned to the cSVD-score if moderate to extensive PVS (score 1-3) were present. These points combined could yield a maximum cSVD-score of 4 points.

### Statistics

To examine the differences in characteristics between hypertensive patients and controls, Pearson chi-square or Fisher’s exact tests were applied for categorical variables when appropriate. For continuous variables independent Student’s *t*-tests were performed. Outliers were defined as a data point with a deleted studentized residual higher than 3.5, and Cook’s distance higher than 1.

First, we set out to determine the association between *K_i_* or *v_p_* in the ROIs (i.e., total GM, total WM, deep GM, cortical GM) with hypertension status by performing multivariable linear regression analyses, corrected for age and sex. Additionally, multivariable linear regression analyses corrected for age and sex were applied to investigate the relation between the measured blood pressure characteristics – SBP, DBP and MAP – and the *K_i_* and *v_p_* in the brain ROIs. Furthermore, these analyses were repeated while alternately adjusting for relevant cardiovascular risk factors, which are BMI, alcohol use, history of smoking, and hematocrit level. Intake of antihypertensive medication and cholesterol lowering medication was also considered alternately in the regression analyses. Lastly, to study the potential influence of cSVD on the associations, the analyses were adjusted by total cSVD score.

An additional post-hoc analysis was performed to explore the difference in leakage measures between the subgroups of hypertensive patients, which were based on the measured blood pressure on the day of the MRI-scan, and their blood pressure medication intake. This resulted in three subgroups: NT+: patients who had normal blood pressure levels, using antihypertensive medication (n=12), HT-: patients with high blood pressure, without using antihypertensive medication (n=5), and HT+: patients with high blood pressure, despite of using antihypertensive medication (n=5). To explore the between-group differences, ANOVA with post-hoc Tukey tests were performed.

A *P*-value < 0.05 was considered statistically significant. All statistical analyses were conducted using SPSS (version 28.0, Chicago, IL).

## Results

The group characteristics of the hypertensive patients (n = 22 with a mean age of 66 years of whom 64% are female) and the controls (n = 19 with a mean age of 62 years, of whom 42% are female) who were included for the final analysis are listed in Table 1. One control participant and one hypertensive patient were identified as outliers in terms of leakage rates and therefore excluded from further analysis. SBP was found to be significantly higher for the hypertensive patients than for the controls. No significant differences in blood pressure measures, age, sex, cardiovascular risk factors, blood measures, or cSVD scores were found. Examining the association between sex and *K_i_* showed that the leakage rate was significantly higher in males (*P* < 0.001 in all regions), but no significant relation with age was found.

**Table 1:**
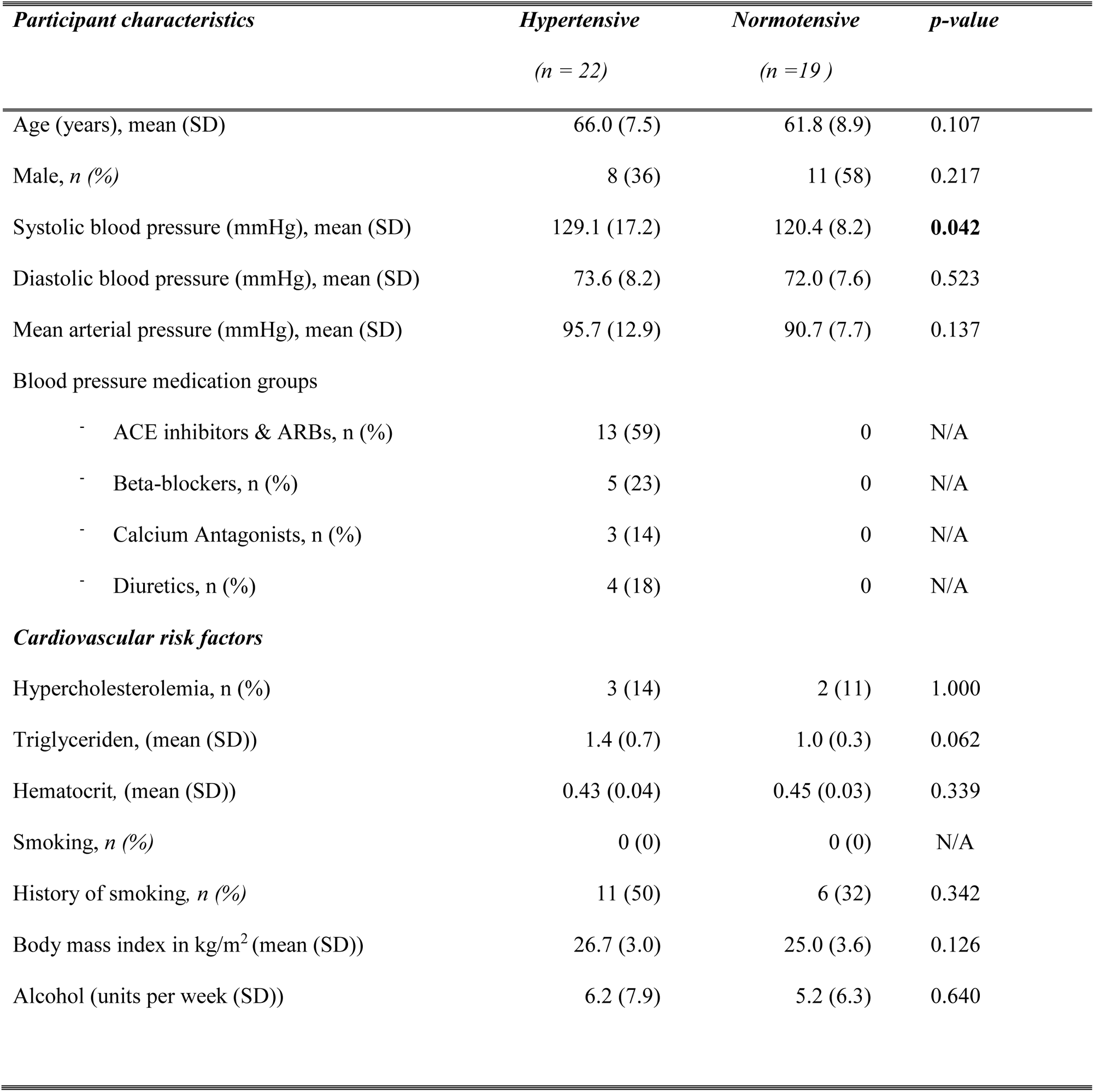
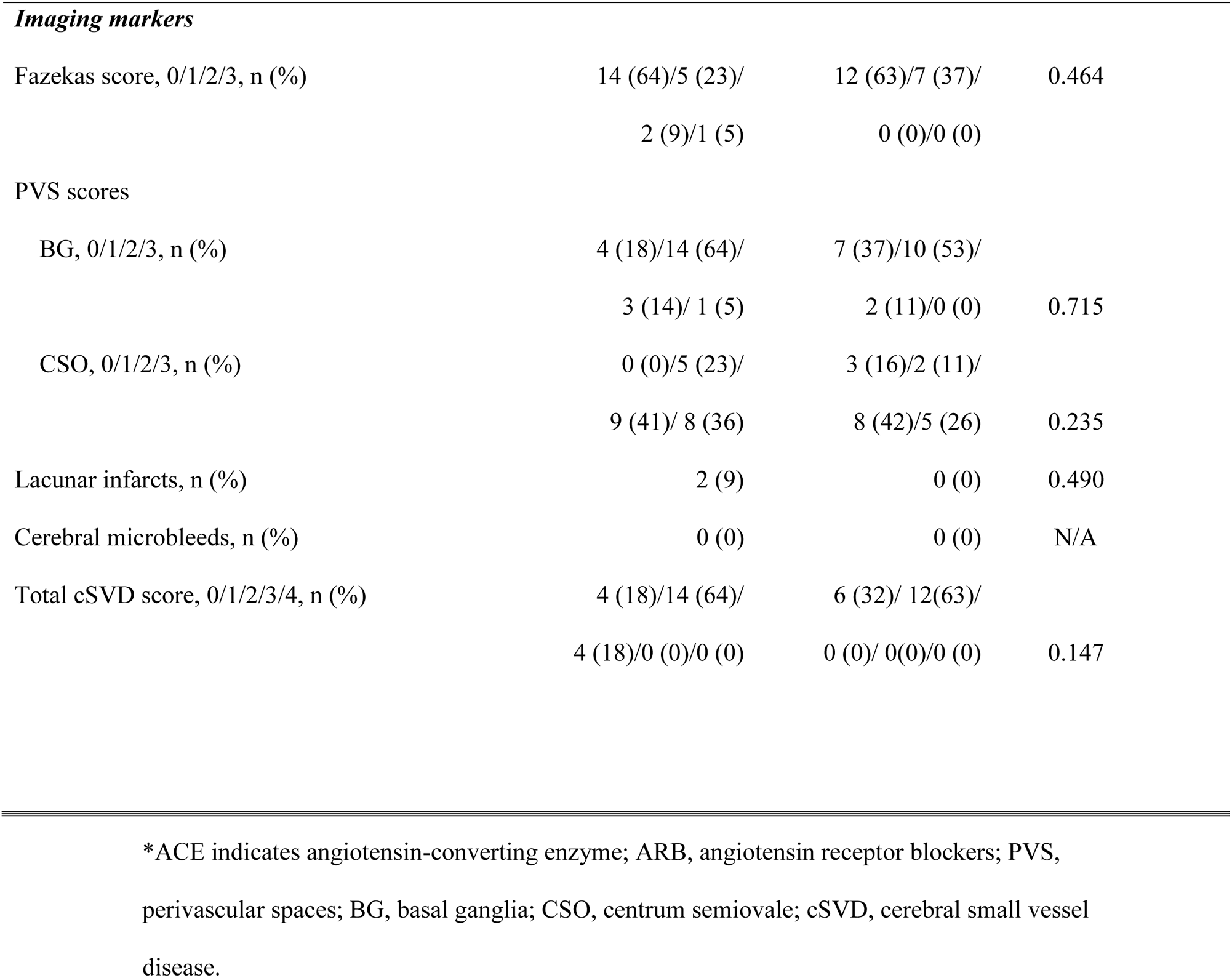
Characteristics of hypertensive patients and normotensive control participants. Significant differences are depicted in bold.

| *Participant characteristics* | *Hypertensive*<br>(*n* = 22) | *Normotensive*<br>(*n* = 19 ) | *p-value* |
| --- | --- | --- | --- |
| Age (years), mean (SD) | 66.0 (7.5) | 61.8 (8.9) | 0.107 |
| Male, *n* (%) | 8 (36) | 11 (58) | 0.217 |
| Systolic blood pressure (mmHg), mean (SD) | 129.1 (17.2) | 120.4 (8.2) | **0.042** |
| Diastolic blood pressure (mmHg), mean (SD) | 73.6 (8.2) | 72.0 (7.6) | 0.523 |
| Mean arterial pressure (mmHg), mean (SD) | 95.7 (12.9) | 90.7 (7.7) | 0.137 |
| Blood pressure medication groups |  |  |  |
| - ACE inhibitors & ARBs, *n* (%) | 13 (59) | 0 | N/A |
| - Beta-blockers, *n* (%) | 5 (23) | 0 | N/A |
| - Calcium Antagonists, *n* (%) | 3 (14) | 0 | N/A |
| - Diuretics, *n* (%) | 4 (18) | 0 | N/A |
| *Cardiovascular risk factors* |  |  |  |
| Hypercholesterolemia, *n* (%) | 3 (14) | 2 (11) | 1.000 |
| Triglyceriden, (mean (SD)) | 1.4 (0.7) | 1.0 (0.3) | 0.062 |
| Hematocrit, (mean (SD)) | 0.43 (0.04) | 0.45 (0.03) | 0.339 |
| Smoking, *n* (%) | 0 (0) | 0 (0) | N/A |
| History of smoking, *n* (%) | 11 (50) | 6 (32) | 0.342 |
| Body mass index in kg/m<sup>2</sup> (mean (SD)) | 26.7 (3.0) | 25.0 (3.6) | 0.126 |
| Alcohol (units per week (SD)) | 6.2 (7.9) | 5.2 (6.3) | 0.640 |
| *Imaging markers* |  |  |  |
| Fazekas score, 0/1/2/3, n (%) | 14 (64)/5 (23)/<br>2 (9)/1 (5) | 12 (63)/7 (37)/<br>0 (0)/0 (0) | 0.464 |
| PVS scores |  |  |  |
| BG, 0/1/2/3, n (%) | 4 (18)/14 (64)/<br>3 (14)/ 1 (5) | 7 (37)/10 (53)/<br>2 (11)/0 (0) | 0.715 |
| CSO, 0/1/2/3, n (%) | 0 (0)/5 (23)/<br>9 (41)/ 8 (36) | 3 (16)/2 (11)/<br>8 (42)/5 (26) | 0.235 |
| Lacunar infarcts, n (%) | 2 (9) | 0 (0) | 0.490 |
| Cerebral microbleeds, n (%) | 0 (0) | 0 (0) | N/A |
| Total cSVD score, 0/1/2/3/4, n (%) | 4 (18)/14 (64)/<br>4 (18)/0 (0)/0 (0) | 6 (32)/ 12(63)/<br>0 (0)/ 0(0)/0 (0) | 0.147 |

\*ACE indicates angiotensin-converting enzyme; ARB, angiotensin receptor blockers; PVS, perivascular spaces; BG, basal ganglia; CSO, centrum semiovale; cSVD, cerebral small vessel disease.

The *K_i_* in hypertensive patients did not significantly differ from the *K_i_* in healthy controls, while adjusting for age and sex (Table 2). In contrast, *v_p_* was significantly higher for hypertensive patients in all GM ROIs and showed a comparable trend for the entire WM. Positive significant relations of *K_i_* with systolic and DBP and MAP were obtained in the cortical GM and for DBP and MAP in total GM (Table 3). *K_i_* versus SBP yielded a positive trend towards significance in total GM. No significant relations were found for SBP, DBP and MAP with *K_i_* in WM and deep GM. For *v_p,_* no significant relations were found with blood pressure measures.

**Table 2:** Comparison of *K_i_* and *v_p_* between hypertensive patients and normative control participants, obtained by regression analysis adjusted for age and sex. Significant differences are depicted in bold. *BBB indicates blood-brain barrier; MAP, mean arterial pressure; *K_i_*, blood-brain barrier leakage; *v_p_*, blood plasma volume fraction; WM, white matter; GM, grey matter.

| *Brain region* | $K_i (*10^{-4})$ | | | | $v_p (*10^{-2})$ | | | |
| --- | --- | --- | --- | --- | --- | --- | --- | --- |
|  | Hypertensive | Normotensive | Standardized |  | Hypertensive | Normotensive | Standardized |  |
| | | | $\beta$ | p-value | | | $\beta$ | p-value |
| Total WM | 4.23 ± 1.98 | 4.61 ± 1.64 | 0.026 | 0.847 | 3.12 ± 0.93 | 2.98 ± 0.80 | 0.216 | 0.095 |
| Total GM | 6.59 ± 1.85 | 7.08 ± 1.75 | -0.016 | 0.900 | 4.27 ± 0.93 | 4.04 ± 0.87 | 0.257 | **0.049** |
| Cortical GM | 6.58 ± 1.83 | 7.05 ± 1.69 | -0.010 | 0.940 | 4.23 ± 0.93 | 4.01 ± 0.84 | 0.263 | **0.042** |
| Deep GM | 3.61 ± 2.42 | 3.95 ± 2.00 | 0.043 | 0.683 | 3.30 ± 1.05 | 2.95 ± 1.03 | 0.276 | **0.031** |

**Table 3:** Multivariable linear regression analyses were performed to obtain the association between BBB leakage rate and blood plasma volume fraction, and blood pressure measures, adjusted for age and sex. Significant differences are depicted in bold.

|  | Systolic blood pressure |  | Diastolic blood pressure |  | Mean arterial pressure |  |
| --- | --- | --- | --- | --- | --- | --- |
| **Brain region** | Standardized $\beta$ | p-value | Standardized $\beta$ | p-value | Standardized $\beta$ | p-value |
| <u>$K_i$</u> |  |  |  |  |  |  |
| Total WM | 0.049 | 0.705 | 0.168 | 0.199 | 0.102 | 0.426 |
| Total GM | 0.227 | 0.052 | 0.253 | **0.035** | 0.271 | **0.019** |
| Cortical GM | 0.246 | **0.043** | 0.271 | **0.031** | 0.291 | **0.015** |
| Deep GM | 0.098 | 0.326 | 0.144 | 0.159 | 0.130 | 0.188 |
| <u>$v_p$</u> |  |  |  |  |  |  |
| Total WM | 0.076 | 0.546 | -0.094 | 0.468 | -0.001 | 0.995 |
| Total GM | -0.061 | 0.632 | -0.245 | 0.058 | -0.143 | 0.260 |
| Cortical GM | -0.061 | 0.632 | -0.250 | 0.051 | -0.148 | 0.239 |
| Deep GM | -0.033 | 0.796 | -0.152 | 0.238 | -0.085 | 0.498 |

\*MAP indicates mean arterial pressure; $K_i$ , blood-brain barrier leakage; $v_p$ , blood plasma volume fraction; WM, white matter; GM, grey matter

The scatterplots in Figure 1 show the relation between *K_i_* and MAP in the four ROIs in more detail. Supplemental Figure 1 illustrates the relation between *v_p_* and MAP in the four ROIs.

**Figure 1:**
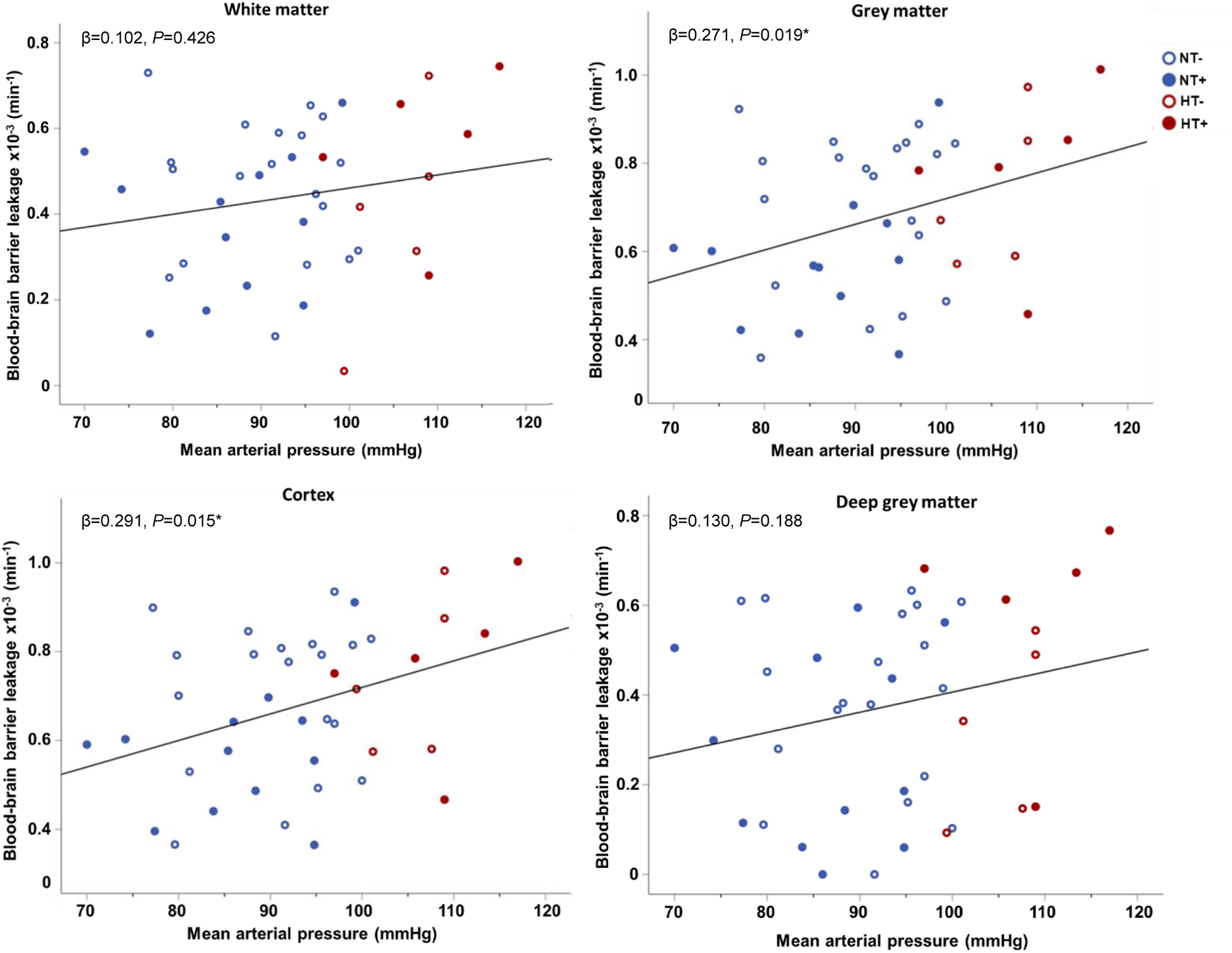
Scatterplots of the blood-brain barrier leakage rate in the four regions of interest versus the mean arterial pressure. The red open and solid data points indicate hypertensive patients without (HT-) and with intake of antihypertensive medication (HT+), respectively, and the blue open and solid data points indicate normotensive participants without (NT-) and with intake of antihypertensive medication (NT+), respectively. * indicates significant associations. Note that the regression line aims to improve visualization, as it is not corrected for age and sex.

**Figure 2.**
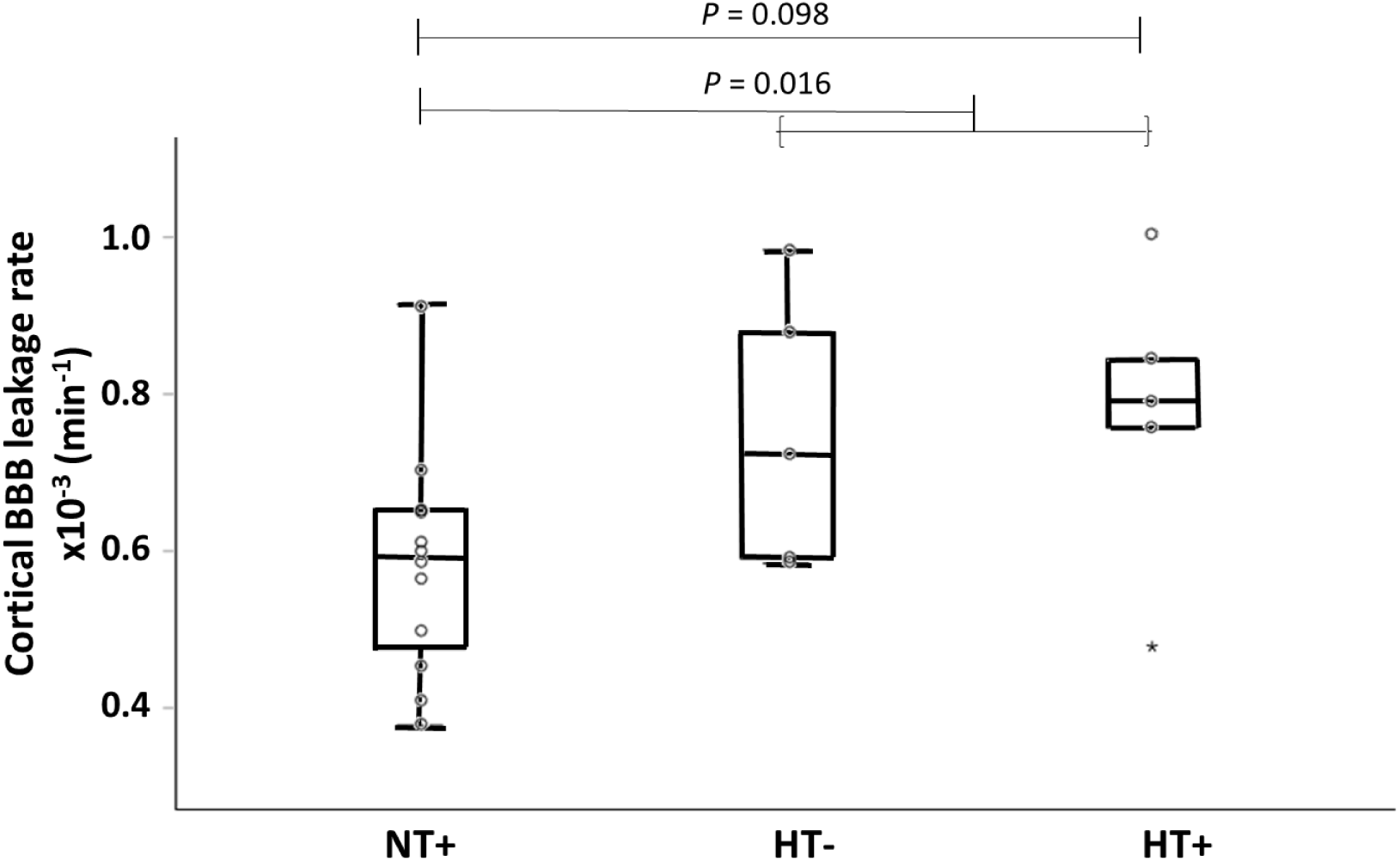
Blood-brain barrier (BBB) leakage in the cortex for the three hypertensive patient subgroups. NT+ indicates normotensive participants with intake of antihypertensive medication, HT- and HT+ hypertensive patients without and with intake of antihypertensive medication, respectively. A trend can be observed between the NT+ and the HT+ group. Note that the combined HT- and HT+ group shows a significant difference with NT+ group.

### Cardiovascular risk factors

The obtained relations between *K_i_* and DBP, and *K_i_* and MAP remained significant after alternately adjusting for cardiovascular risk factors, medication intake and cSVD score. When considering BMI, alcohol intake, and cSVD score in the regression analyses with SBP in the cortical GM, the adjusted relations did not remain significant, but still showed a positive trend towards significance (β = 0.216, *P* = 0.062, β = 0.245, *P* = 0.053 and β = 0.245, *P =* 0.059, respectively). For total GM, the relation was found to be significant when correcting for history of smoking or intake of cholesterol medication (β = 0.231, *P =* 0.050 and β = 0.233, *P =* 0.049, respectively).

### Subgroup analyses

Post-hoc analysis of the hypertensive subgroups (HT+, HT-, and NT+) revealed a trend towards significance of higher *K_i_* in HT+ compared to NT+ patients for the grey matter regions (cortex: *P* = 0.098, total GM: *P* = 0.088, deep GM: *P* = 0.058), but not for WM. After pooling of the HT- and HT+ patients, the NT+ group demonstrated lower leakage rates than the combined HT group for the total and cortical GM (*P* = 0.021 and *P* = 0.016, respectively). Figure 3 shows the corresponding boxplot for the cortex as an example. Boxplots for *K_i_* in the total GM, deep GM and total WM can be found in Supplemental Figure 2. Post-hoc analysis for *v_p_* did not show significant differences between subgroups.

## Discussion

In this study, we set out to investigate the relationship between hypertension, SBP, DBP and MAP, and BBB leakage measured with a highly sensitive DCE-protocol at 7 Tesla MRI in human subjects. Based on dichotomous hypertension status, we found no relation with BBB leakage. For the individual blood pressure characteristics, i.e., SBP, DBP and MAP, a higher blood pressure was associated with stronger BBB leakage in the grey matter and not in the white matter. The results were independent of cardiovascular risk factors and presence of cSVD imaging markers. Medically well treated patients (NT+) showed less BBB leakage than patients with current hypertension (pooled HT- and HT+ group).

The BBB is often assumed to become impaired due to the pathophysiological pathway as a consequence of hypertension.^2,6^ To investigate whether increased blood pressure impacts the BBB integrity prior to the presence of overt cerebral tissue damage, this research focused on differences in BBB leakage between hypertensive patients with no known cerebrovascular disease and normotensive controls. The lack of differences in BBB leakage found between the hypertensive and normotensive group, is likely due to the similarities of the blood pressure characteristics of the two groups. SBP was the only blood pressure measure found to be higher in the hypertensive patients. In addition to the groups being matched beforehand on age and sex, the other cardiovascular risk factors, blood measures, and cSVD scores were also found to be similar across the two groups. However, we observed a large variation in BBB leakage in the hypertensive patients (NT+, HT-, and HT+), which also could explain the absence of a difference in BBB leakage between the hypertension versus normotension group. This demonstrates a large influence of current blood pressure (as opposed to hypertension status) on the BBB integrity. The observed relation between current blood pressure and BBB leakage suggests that a higher blood pressure could lead to endothelial dysfunction, impairing the permeability of the barrier.

In the GM, a positive relation between blood pressure and BBB leakage was obtained. In contrast, no relation was obtained within the WM. Our findings imply that in grey matter, especially in the well perfused cortical grey matter, subtle leakage is easier, and therefore earlier, to detect than in the less perfused white matter. Additionally, the tissue around the basal ganglia, thus WM, is more protected from the effects of cerebrovascular dysfunction as these arterioles contain two layers, whereas the superficial perforating arterioles have only one layer.^20^

Previous studies using hypertension models in animals showed that a higher BBB permeability is related to hypertension^6,21,22^, although some studies did not find any differences in hypertensive animal models.^23,24^ It should be noted that these studies use different methods of measuring BBB leakage, as they use other contrast agents, such as fluorescently labeled dextran or lectin.^21,22,25^ Furthermore, these studies are able to dissect the brain after imaging, to investigate the neurovascular properties in more detail. Studies on BBB leakage in humans often focus on diagnosed cerebrovascular diseases, such as cSVD and (mixed) dementia.^8,9,26,27^ These studies demonstrated stronger BBB leakage in these patients, and showed that hypertension is a significant covariate. However, they did not investigate hypertension as a separate condition, or in subjects without overt neurovascular disease.

The associations with blood plasma volume fraction were contrary to the findings of the leakage rate in this study. Differences in blood plasma volume fraction were found in hypertensive patients versus normotensive controls, based on dichotomous status, while no relations were found for the current blood pressure measures. These results are contradictory to the pathway of a reduced cerebral blood flow, which is related to the blood plasma volume fraction, in hypertension. However, the higher blood plasma volume in hypertensive patients is not indicative of an advanced cerebrovascular disease stage, as vessels tend to narrow (lower blood volume or flow) in a more advanced hypertensive disease state.^28^

As previous studies showed an association between age and sex, and BBB leakage, we initially adjusted for these two covariates.^31–33^ We indeed found a strong correlation between sex and leakage measures, indicating that women have a lower BBB permeability compared to men. This effect could be explained by the hormonal differences between men and women, as female hormones, such as estrogen, may act neuroprotective.^32,33^

### Cardiovascular risk factors

This study assessed several cardiovascular risk factors, as well as a cSVD score which was rated on a scale based on previous literature and reflects the total cSVD burden.^14,29^ All observed associations remained significant after adjusting for these covariates. In contrast to our findings, previous studies showed that cSVD was associated with higher BBB leakage.^8,30^ The low number of cSVD markers observed in our population compared to the more severe cSVD rates in other studies could account for this divergence. We therefore emphasize that in this study, we set out to measure this specific consequence of microvascular damage in a rather early stage of (or mild) cSVD.

### Hypertension subgroups

As mentioned above, our results show a large effect on the BBB leakage rate of the actual blood pressure, measured preceding the MRI examination, rather than hypertension status. While comparing the leakage rates between the hypertensive subgroups, a trend of stronger BBB leakage was found in hypertensive patients with inadequate antihypertensive medication (HT+), compared to patients with effective antihypertensive medication (NT+). The HT-group tends to be associated with higher BBB leakage values than the NT+ group, but only slightly lower values than the HT+ group. As the HT+ group does not show significantly higher or lower values than the HT-group, and the combined HT group obtained higher leakage values than the NT+ group, we suggest that effective medication has a protective effect on BBB permeability. Previous studies also have shown such restoring effects of specific antihypertensive medication in hypertensive rat models.^6^ However, it is important to be aware of the small sample size of the subgroups.

### Strengths and limitations

This study focused on BBB leakage in patients with high blood pressure, without known cerebrovascular diseases, compared to normotensive controls. The strength of this study design is that it enabled the investigation of early BBB changes, before neurovascular damage is visible on the MRI scans and before subjects experience neurological complaints.

Furthermore, a sparsely sampled, interleaved MRI protocol was used to obtain subtle BBB measures, which shortened the scanning time substantially compared to frequently used DCE sequences lasting up to 30 minutes. This protocol also allows for other imaging sequences to be scanned in between, allowing to use the scanning time more efficient. Furthermore, using high-field 7T MRI enhances the signal-to-noise ratio with increasing spatial resolution, thereby enabling improved measurement of subtle BBB leakage.

There are several methodological considerations. The blood pressure was measured using a 30-minute during automatic protocol, although the most accurate method would be 24-hour ambulatory blood pressure monitoring. Regardless, the blood pressure measurement protocol in this study is validated to be the second most accurate method.^34^

As discussed, the cSVD scores were relatively low in our population. Future studies should also include participants with higher cSVD scores to improve the ability of investigating BBB leakage in patients with hypertension with varying amounts of visibly developed neurovascular disease.

Larger sample sizes in the three hypertension subgroups, or a longitudinal study design (before and after start of antihypertensive treatment) would also be recommended in future research that focuses on studying medication effects on BBB leakage in more detail. While previous studies show varying effects between the different types of antihypertensive medication, our study did not have enough power in the subgroups to draw definite conclusions, as our study was not designed for this specific aim.

### Perspectives

To summarize, this study aimed to investigate the relationship between BBB integrity, high blood pressure and hypertension. For the dichotomous hypertension status, no significant effects of BBB leakage were found. However, when looking at continuous blood pressure measures, a strong positive relation was found between leakage in the cortical grey matter and current blood pressure. A protective function of antihypertensive medication was observed, but future studies should include more hypertensive patients or engage a longitudinal study design to investigate this in more detail.

## Non-standard Abbreviations and Acronyms

BBB: blood-brain barrier
cSVD: cerebral small vessel disease
DBP: diastolic blood pressure
DCE-MRI: dynamic contrast enhanced magnetic resonance imaging
GM: grey matter
MAP: mean arterial pressure
PVS: perivascular spaces
ROI: region of interest
SBP: systolic blood pressure
TE: echo time
TR: repetition time
VIF: vascular input function
WM: white matter
WMH: white matter hyperintensities

## Acknowledgements

We thank the technicians of Scannexus for assisting with scanning and the administration of the contrast agent.

## Sources of Funding

This study was partly funded by the EU’s Horizon 2020 project ‘CRUCIAL’, Grant agreement number 848109, and ‘Stichting De Weijerhorst Foundation’.

## Disclosures

None

## Supplemental Material

Figure S1-S2

Tables S1

## Notes

### Competing Interest Statement

The authors have declared no competing interest.

### Clinical Trial

https://trialsearch.who.int/Trial2.aspx?TrialID=NL7537

### Author Declarations

Medical Ethical Committee of Maastricht University Medical Center

## References

1. Mills KT, Stefanescu A, He J. The global epidemiology of hypertension. Nat. Rev. Nephrol. 2020;16:223–237.

2. Faraco G, Iadecola C. Hypertension: A Harbinger of Stroke and Dementia. Hypertension. 2013;62:810–817.

3. Iadecola C, Yaffe K, Biller J, Bratzke LC, Faraci FM, Gorelick PB, Gulati M, Kamel H, Knopman DS, Launer LJ, et al. Impact of Hypertension on Cognitive Function: A Scientific Statement From the American Heart Association. Hypertension. 2016;68.

4. Meissner A. Hypertension and the Brain: A Risk Factor for More Than Heart Disease. Cerebrovasc. Dis. 2016;42:255–262.

5. Sweeney MD, Zhao Z, Montagne A, Nelson AR, Zlokovic BV. Blood-Brain Barrier: From Physiology to Disease and Back. Physiol. Rev. 2019;99:21–78.

6. Katsi V, Marketou M, Maragkoudakis S, Didagelos M, Charalambous G, Parthenakis F, Tsioufis C, Tousoulis D. Blood–brain barrier dysfunction: the undervalued frontier of hypertension. J. Hum. Hypertens. 2020;34:682–691.

7. Canjels LPW, Jansen JFA, Alers RJ, Ghossein-Doha C, Van Den Kerkhof M, Schiffer VMMM, Mulder E, Gerretsen SC, Aldenkamp AP, Hurks PPM, et al. Blood–brain barrier leakage years after pre-eclampsia: dynamic contrast-enhanced 7-TESLA MRI study. Ultrasound Obstet. Gynecol. 2022;60:541–548.

8. Thrippleton MJ, Backes WH, Sourbron S, Ingrisch M, van Osch MJP, Dichgans M, Fazekas F, Ropele S, Frayne R, van Oostenbrugge RJ, et al. Quantifying blood-brain barrier leakage in small vessel disease: Review and consensus recommendations. Alzheimers Dement. 2019;15:840–858.

9. Dobrynina LA, Shamtieva KV, Kremneva EI, Zabitova MR, Akhmetzyanov BM, Gnedovskaya EV, Krotenkova MV. Daily blood pressure profile and blood–brain barrier permeability in patients with cerebral small vessel disease. Sci. Rep. 2022;12:7723.

10. Raja R, Rosenberg GA, Caprihan A. MRI measurements of Blood-Brain Barrier function in dementia: A review of recent studies. Neuropharmacology. 2018;134:259–271.

11. Kerkhof M, Voorter PHM, Canjels LPW, Jong JJA, Oostenbrugge RJ, Kroon AA, Jansen JFA, Backes WH. Time-efficient measurement of subtle blood–brain barrier leakage using a T _1_ mapping MRI protocol at 7 T. Magn. Reson. Med. 2021;85:2761–2770.

12. Fischl B, Salat DH, Busa E, Albert M, Dieterich M, Haselgrove C, van der Kouwe A, Killiany R, Kennedy D, Klaveness S, et al. Whole Brain Segmentation. Neuron. 2002;33:341–355.

13. Patlak CS, Blasberg RG, Fenstermacher JD. Graphical Evaluation of Blood-to-Brain Transfer Constants from Multiple-Time Uptake Data. J. Cereb. Blood Flow Metab. 1983;3:1–7.

14. Staals J, Makin SDJ, Doubal FN, Dennis MS, Wardlaw JM. Stroke subtype, vascular risk factors, and total MRI brain small-vessel disease burden. Neurology. 2014;83:1228–1234.

15. Fazekas F, Chawluk J, Alavi A, Hurtig H, Zimmerman R. MR signal abnormalities at 1.5 T in Alzheimer’s dementia and normal aging. Am. J. Roentgenol. 1987;149:351–356.

16. Klarenbeek P, van Oostenbrugge RJ, Rouhl RPW, Knottnerus ILH, Staals J. Ambulatory Blood Pressure in Patients With Lacunar Stroke: Association With Total MRI Burden of Cerebral Small Vessel Disease. Stroke. 2013;44:2995–2999.

17. Wardlaw JM, Benveniste H, Nedergaard M, Zlokovic BV, Mestre H, Lee H, Doubal FN, Brown R, Ramirez J, MacIntosh BJ, et al. Perivascular spaces in the brain: anatomy, physiology and pathology. Nat. Rev. Neurol. 2020;16:137–153.

18. Francis F, Ballerini L, Wardlaw JM. Perivascular spaces and their associations with risk factors, clinical disorders and neuroimaging features: A systematic review and meta-analysis. Int. J. Stroke. 2019;14:359–371.

19. Kerkhof M van den, Thiel MM van der, Oostenbrugge RJ van, Postma AA, Kroon AA, Backes WH, Jansen JF. Impaired damping of cerebral blood flow velocity pulsatility is associated with the number of perivascular spaces as measured with 7T MRI. J. Cereb. Blood Flow Metab. 0:0271678X231153374.

20. Wardlaw JM, Smith C, Dichgans M. Mechanisms of sporadic cerebral small vessel disease: insights from neuroimaging. Lancet Neurol. 2013;12:483–497.

21. Santisteban MM, Iadecola C, Carnevale D. Hypertension, Neurovascular Dysfunction, and Cognitive Impairment. Hypertension. 2023;80:22–34.

22. Biancardi VC, Son SJ, Ahmadi S, Filosa JA, Stern JE. Circulating Angiotensin II Gains Access to the Hypothalamus and Brain Stem During Hypertension via Breakdown of the Blood–Brain Barrier. Hypertension. 2014;63:572–579.

23. Naessens DMP, De Vos J, VanBavel E, Bakker ENTP. Blood–brain and blood– cerebrospinal fluid barrier permeability in spontaneously hypertensive rats. Fluids Barriers CNS. 2018;15:26.

24. Rodrigues SF, Granger DN. Cerebral Microvascular Inflammation in DOCA Salt-Induced Hypertension: Role of Angiotensin II and Mitochondrial Superoxide. J. Cereb. Blood Flow Metab. 2012;32:368–375.

25. Santisteban MM, Ahn SJ, Lane D, Faraco G, Garcia-Bonilla L, Racchumi G, Poon C, Schaeffer S, Segarra SG, Körbelin J, et al. Endothelium-Macrophage Crosstalk Mediates Blood-Brain Barrier Dysfunction in Hypertension. Hypertension. 2020;76:795–807.

26. Chagnot A, Barnes SR, Montagne A. Magnetic Resonance Imaging of Blood–Brain Barrier permeability in Dementia. Neuroscience. 2021;474:14–29.

27. Sweeney MD, Sagare AP, Zlokovic BV. Blood–brain barrier breakdown in Alzheimer disease and other neurodegenerative disorders. Nat. Rev. Neurol. 2018;14:133–150.

28. Iadecola C, Gottesman RF. Neurovascular and Cognitive Dysfunction in Hypertension: Epidemiology, Pathobiology, and Treatment. Circ. Res. 2019;124:1025–1044.

29. Wardlaw JM, Smith EE, Biessels GJ, Cordonnier C, Fazekas F, Frayne R, Lindley RI, O’Brien JT, Barkhof F, Benavente OR, et al. Neuroimaging standards for research into small vessel disease and its contribution to ageing and neurodegeneration. Lancet Neurol. 2013;12:822–838.

30. Wong SM, Jansen JFA, Zhang CE, Staals J, Hofman PAM, van Oostenbrugge RJ, Jeukens CRLPN, Backes WH. Measuring subtle leakage of the blood-brain barrier in cerebrovascular disease with DCE-MRI: Test-retest reproducibility and its influencing factors: Reproducibility of DCE-MRI in Subtle Leakage. J. Magn. Reson. Imaging. 2017;46:159–166.

31. Verheggen ICM, De Jong JJA, Van Boxtel MPJ, Gronenschild EHBM, Palm WM, Postma AA, Jansen JFA, Verhey FRJ, Backes WH. Increase in blood–brain barrier leakage in healthy, older adults. GeroScience. 2020;42:1183–1193.

32. Weber CM, Clyne AM. Sex differences in the blood–brain barrier and neurodegenerative diseases. APL Bioeng. 2021;5:011509.

33. Dion-Albert L, Bandeira Binder L, Daigle B, Hong-Minh A, Lebel M, Menard C. Sex differences in the blood–brain barrier: Implications for mental health. Front. Neuroendocrinol. 2022;65:100989.

34. Williams B, Mancia G, Spiering W, Agabiti Rosei E, Azizi M, Burnier M, Clement DL, Coca A, De Simone G, Dominiczak A, et al. 2018 ESC/ESH Guidelines for the management of arterial hypertension. Eur. Heart J. 2018;39:3021–3104.

